# Development of a standards-based city-wide health information exchange for public health in response to COVID-19

**DOI:** 10.1101/2020.08.12.20173559

**Authors:** Bala Hota, Paul Casey, Anne F. McIntyre, Jawad Khan, Shafiq Rab, Aneesh Chopra, Omar Lateef, Jennifer E. Layden

**Affiliations:** Rush University Medical Center, Chicago, IL, 60612; Centers for Disease Control and Prevention, Atlanta, GA; Care Journey; Chicago Department of Public Health, Chicago, IL

**Keywords:** SARS-CoV2, COVID-19, Public Health Informatics

## Abstract

**Background:** Disease surveillance is a critical function of public health, provides essential information about disease burden, clinical and epidemiologic parameters of disease, and is an important element to effective and timely case and contact tracing. The COVID-19 pandemic has demonstrated the essential role these functions have to preserve public health. Syndromic surveillance, electronic laboratory reporting in the meaningful use program, and the growth of the National Healthcare Safety Network (NHSN) have created linkages between hospitals, commercial labs, and public health that can collect and organize data, often through EHR and order workflows, to improve the timeliness and completeness of reporting. In theory, the standard data formats and exchange methods provided by meaningful use should enable rapid healthcare data exchange in the setting of disruptive healthcare events like a pandemic. In reality, access to data remains challenging, and even if available, often lack conformity to regulated standards.

**Objective:** We sought to use regulated interoperability standards already in production to generate regional bed capacity awareness, enhance the capture of epidemiological risk factors and clinical variables among COVID-19 tested patients, and reduce the administrative burden of reporting for stakeholders in a manner that could be replicated by other public health agencies.

**Methods:** Following a public health order mandating data submission, we developed technical infrastructure to combine multiple data feeds from electronic health record systems. We measured the completeness of each feed, and the match rate between feeds.

**Results:** A cloud-based environment was created that received data from electronic lab reporting, consolidated clinical data architecture, and bed capacity data feeds from sites. Data governance was planned from the project beginning to aid in consensus and principles for data use. 88,906 total persons from CCDA data among 14 facilities, and 408,741 persons from ELR records among 88 facilities, were submitted. Fields routinely absent from ELR feeds included travel histories, clinical symptoms, and comorbidities. CCDA data provided an improvement in the quality of data available for surveillance and was highly complete with <5% for all records types with the exception of patient cell phone. 90.1% of records could be matched between CCDA and ELR feeds.

**Conclusions:** We describe the development of a city-wide public health data hub for the surveillance of COVID-19 infection. We were able to assess the completeness of existing ELR feeds, augment these feeds with CCDA documents, establish secure transfer methods for data exchange, develop cloud-based architecture to enable secure data storage and analytics, and produced meaningful dashboards for the monitoring of capacity and disease burden. We see this public health and clinical data registry as an informative example of the power of common standards across electronic records, and a potential template for future extension of the use of standards to improve public health surveillance.

## Introduction

Since the emergence of SARS CoV-2 from Wuhan, China (1), a global pandemic has been declared(2) and widespread, sustained transmission has been observed across the United States. It has at the time of this writing infected 10.9 million and caused over 520,000 deaths worldwide, and over 2.7 million cases and over 120,000 deaths in the United States, with recent increased incidence rates in states in the Southern and Southwestern U.S.

COVID-19 is a reportable disease in Illinois, and both the Illinois Department of Public Health (IDPH) and Chicago Department of Public Health (CDPH) mandated the reporting of all patents tested for the SARS-CoV-2 virus. Disease surveillance is a critical function of public health, and provides essential information about disease burden, clinical and epidemiologic parameters of disease, and is an important element to effective and timely case and contact tracing. In addition to individual and aggregate level patient data, this pandemic has required careful monitoring of healthcare capacity and utilization to ensure clinical care needs be met.

Support for the public health functions of the surveillance and epidemiology of disease has been embedded in key national informatics initiatives for nearly two decades. These efforts have included syndromic surveillance(3), electronic laboratory reporting(4) in the meaningful use program(5), and the growth of the National Healthcare Safety Network (NHSN)(6). These programs have created linkages between hospitals, commercial labs, and public health that can collect and organize data, often through EHR and order workflows, to improve the timeliness and completeness of reporting.

In theory, the standard data formats and exchange methods provided by meaningful use should enable rapid healthcare data exchange in the setting of disruptive healthcare events like a pandemic. In reality, access to data remains challenging, and even if available, often lack conformity to regulated standards(7). The current COVID-19 pandemic has revealed gaps in data liquidity and the resultant difficulty in gathering information quickly(8).

In the early phase of the pandemic, CDPH and health systems aimed to address two major challenges first, the ability to efficiently submit necessary clinical data elements for COVID tested patients,, and second, the ability to capture aggregate capacity data for resource planning in an administratively efficient manner. Despite significant EHR investments among the city’s hospitals and health systems,, the inability for electronic health records systems to automate delivery of important data elements to public health surveillance systems meant that providers and health systems had to manually enter data into the public health reporting system. However, the high volume of patients and significant work demands on health systems limited timely, and complete manual data entry. As the pandemic unfolded, multiple agencies had requested bed and surge capacity information, including NHSN, FEMA, the National guard, the Illinois Department of Public Health, all with slightly varying definitions (Table 1). Locally, an important aspect of capturing the resource capacity data was to monitor surge capacity and assist with coordination of resources. The multiple reporting requirements, varied definitions, and limited mechanisms for automated, real time submission of key resource metrics such as bed capacities raised concern about the ability to locally monitor the resource capacity across our systems.

**Table 1.**
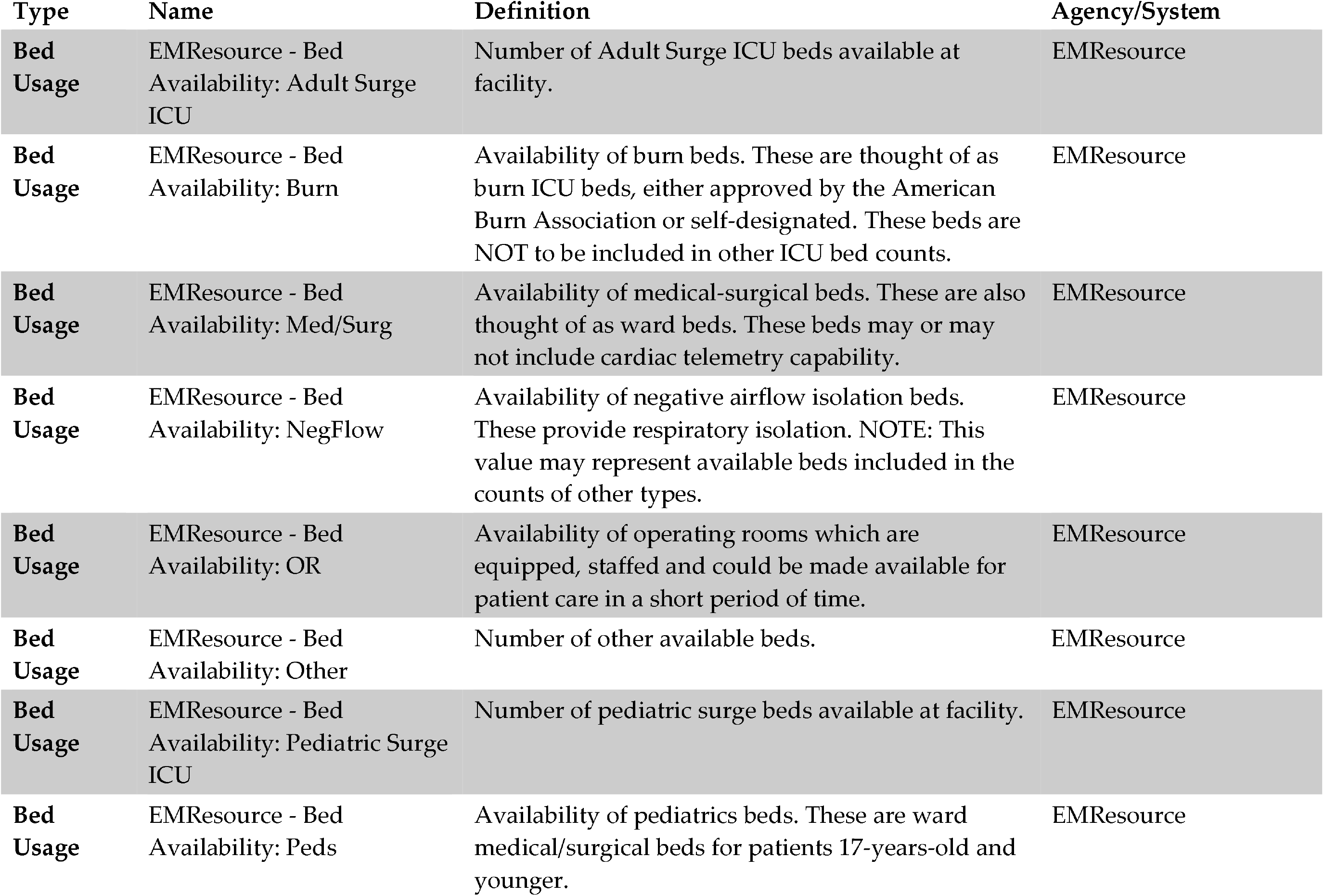
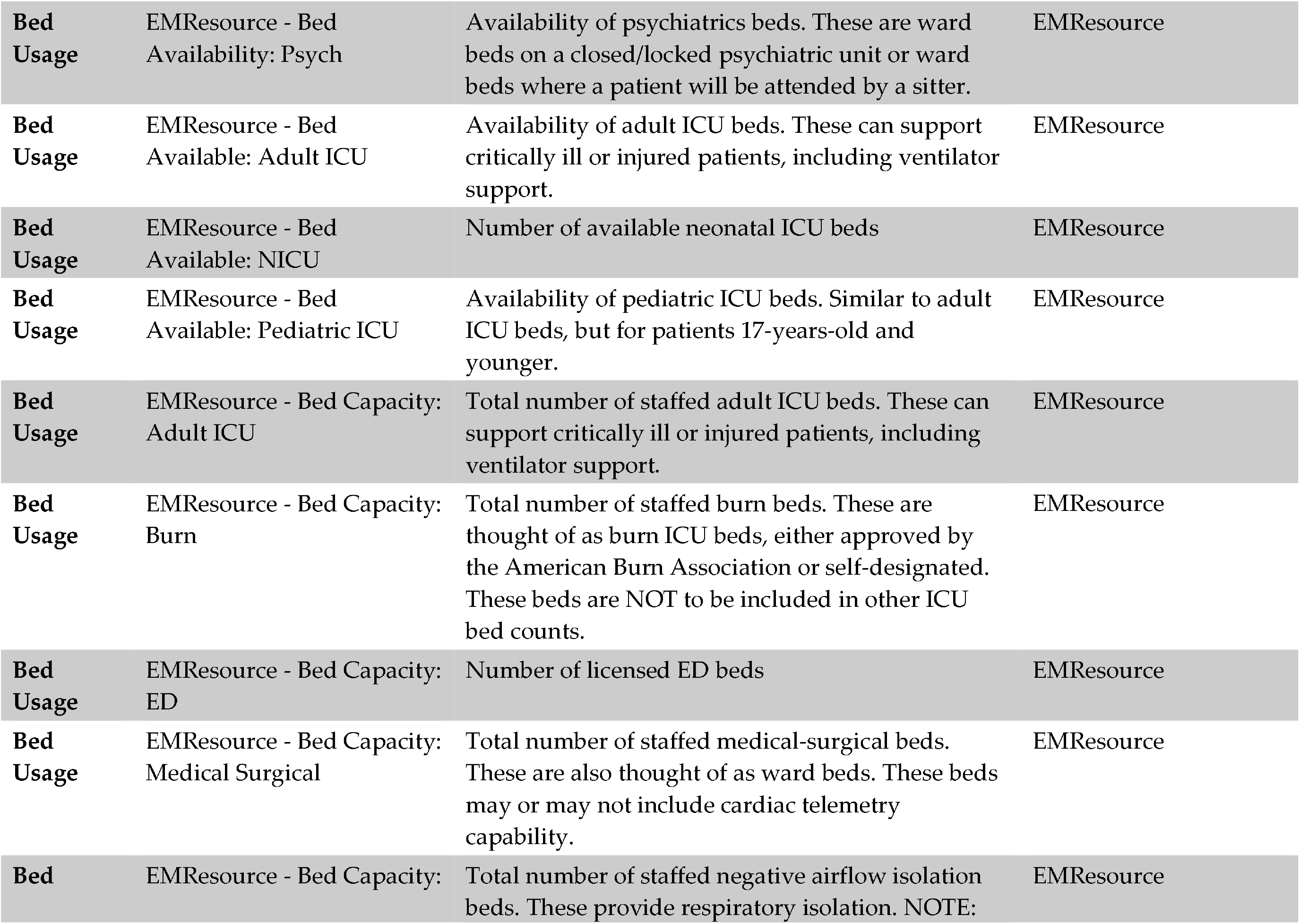
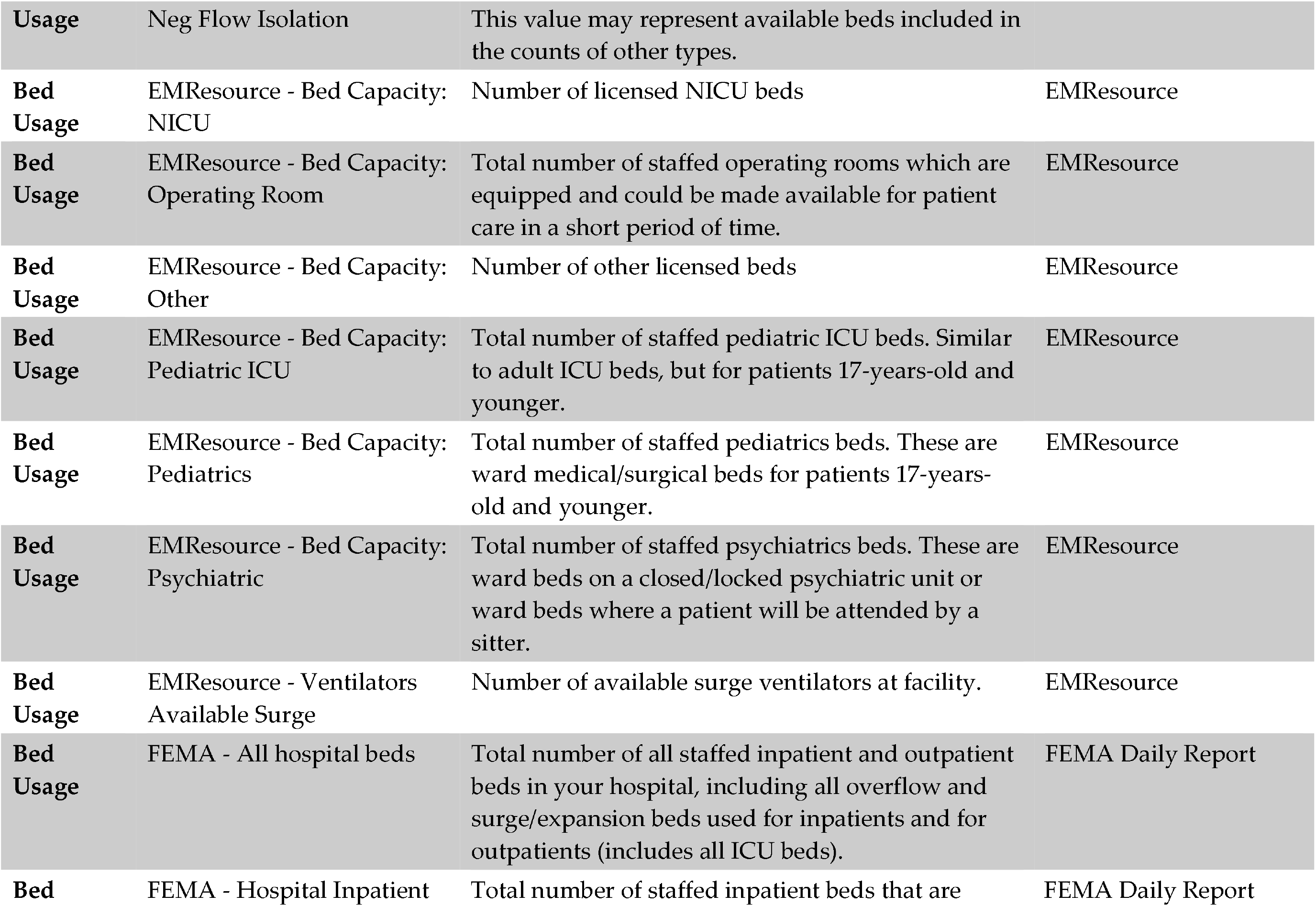
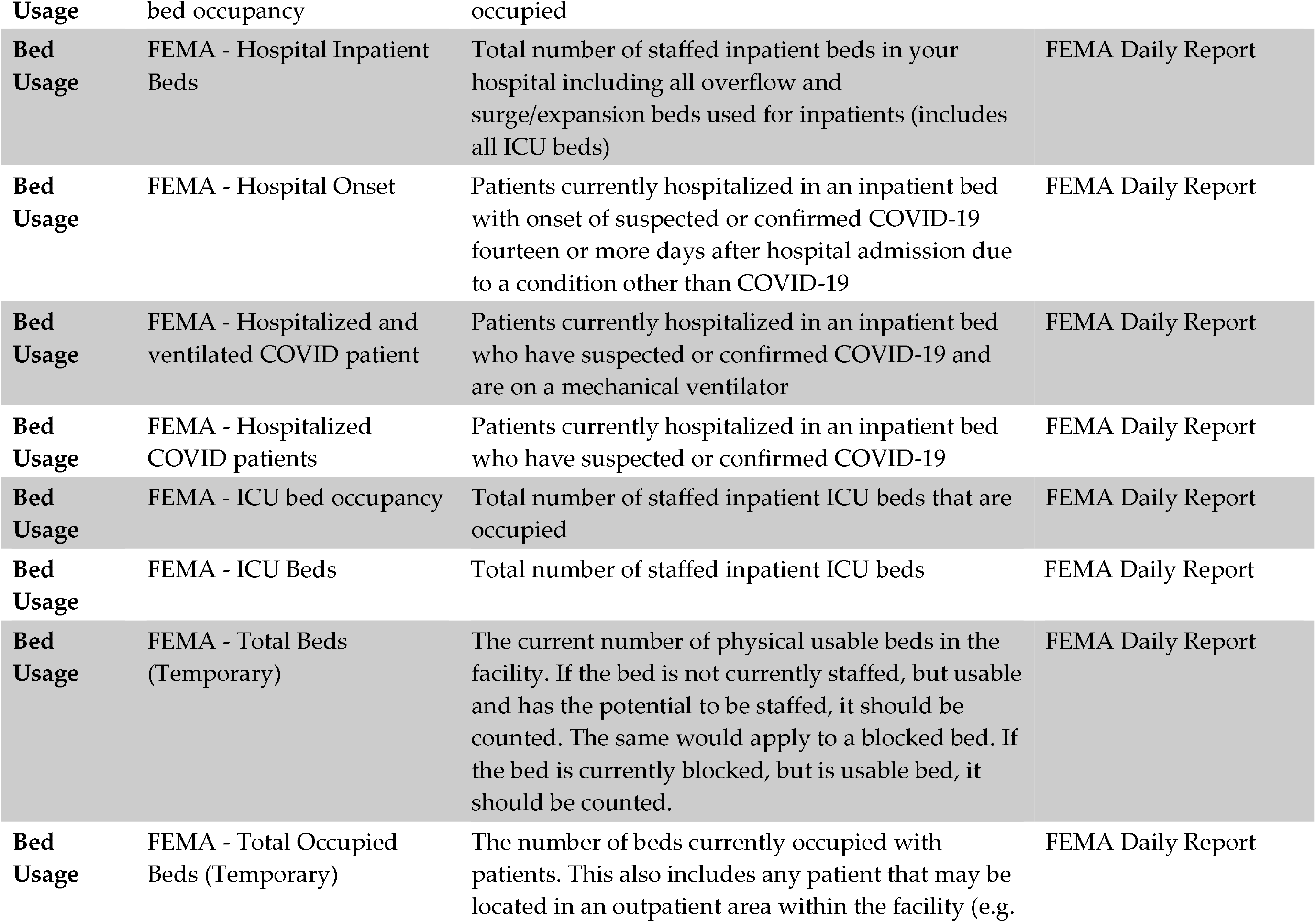
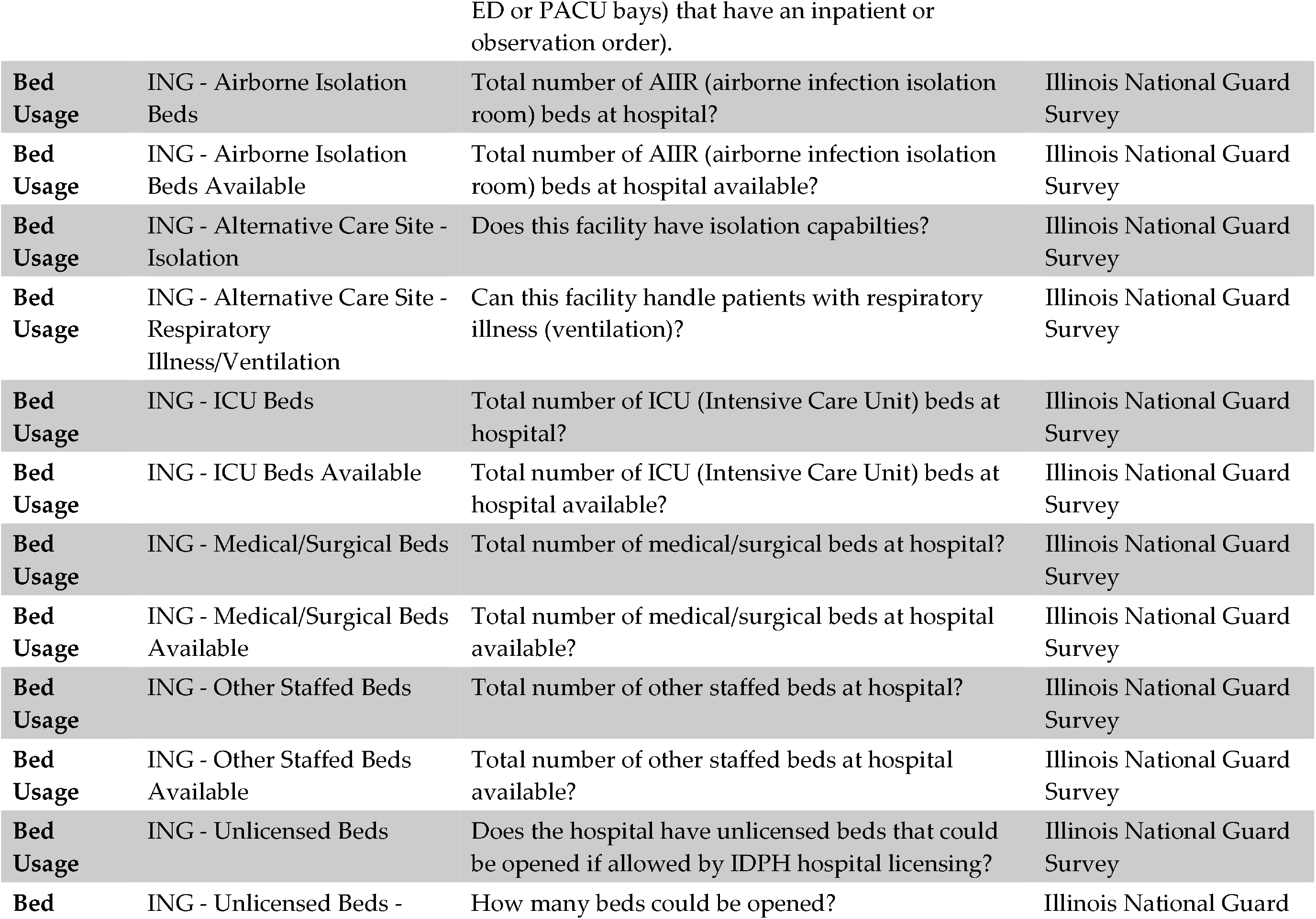
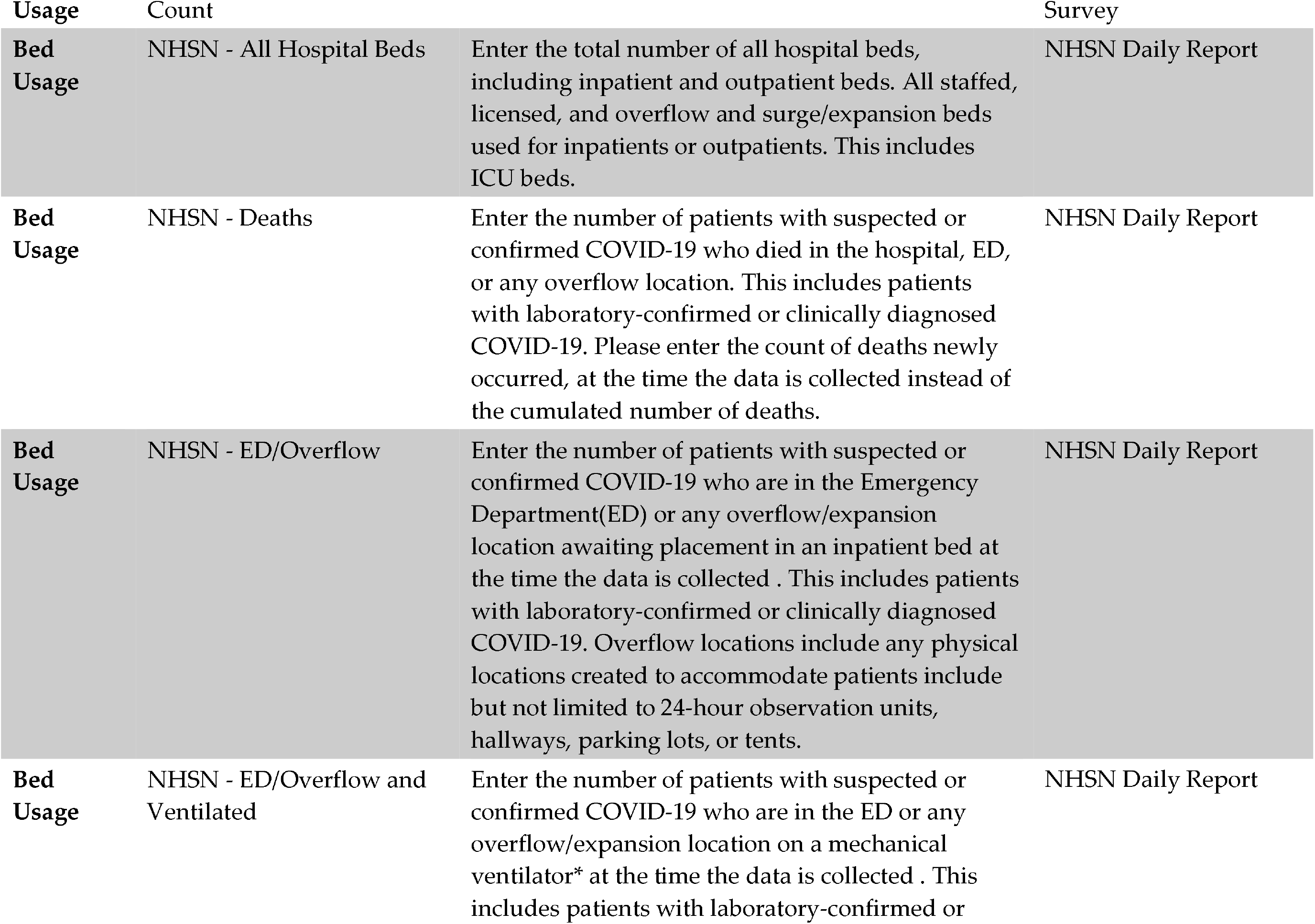
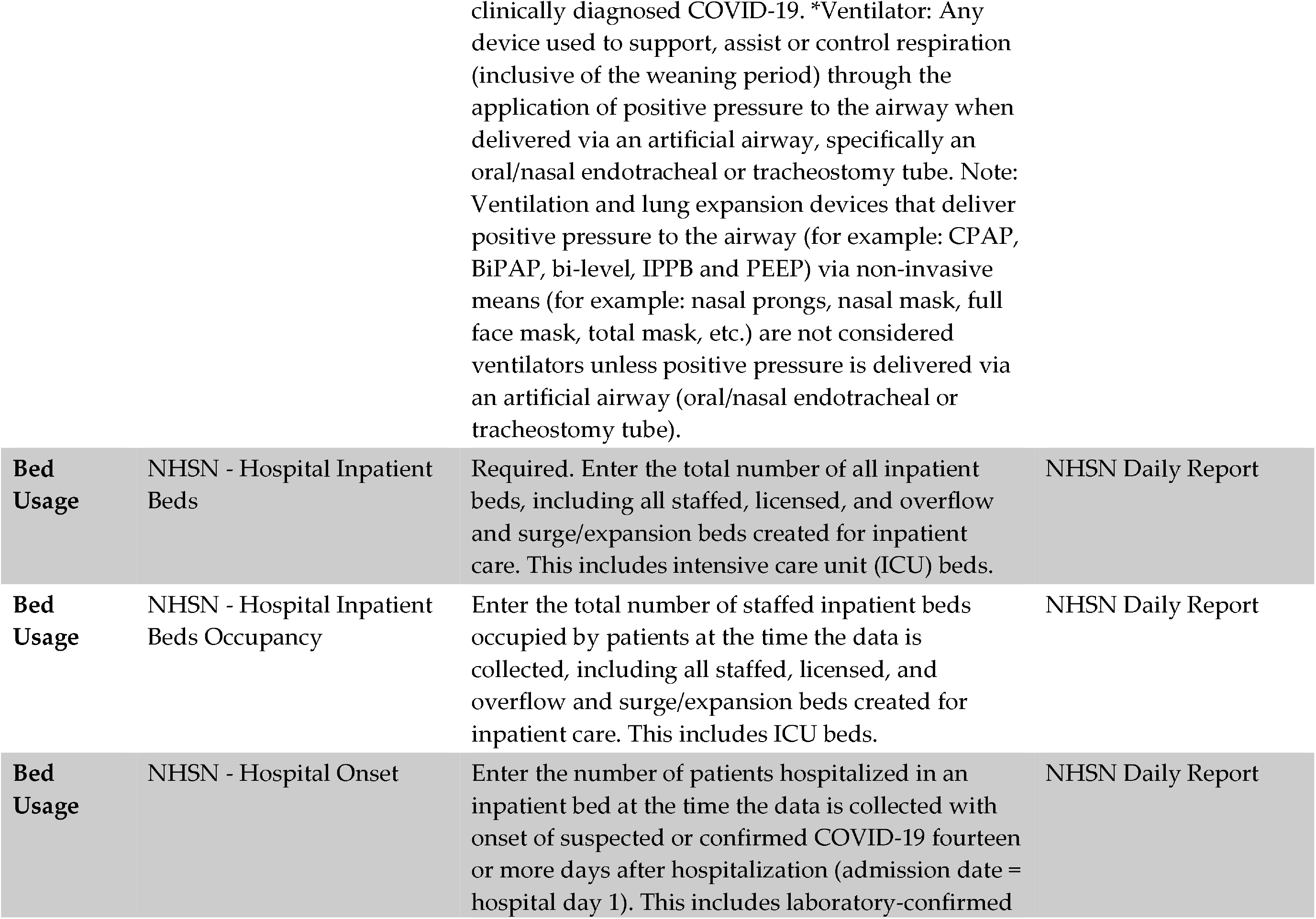
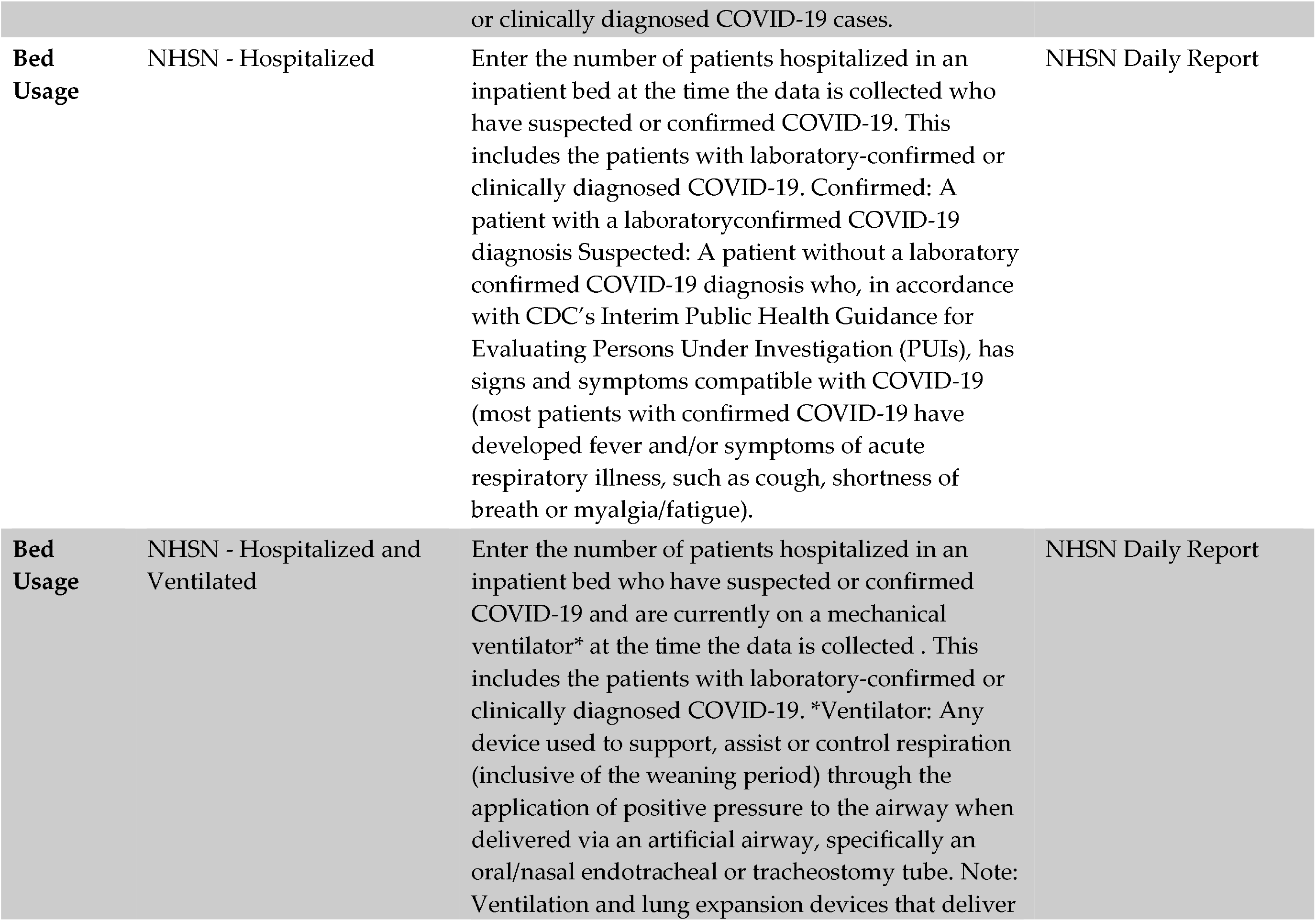
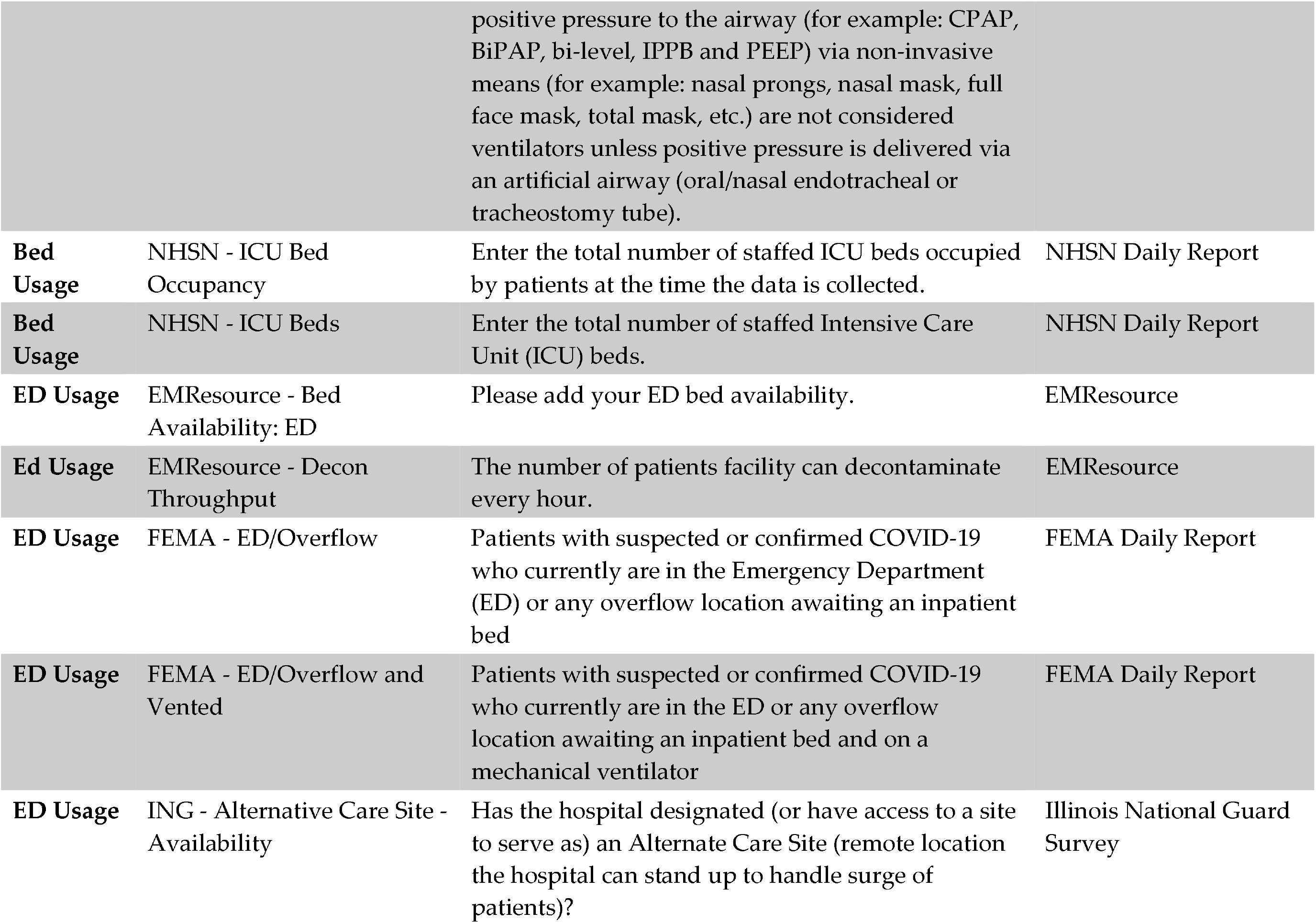
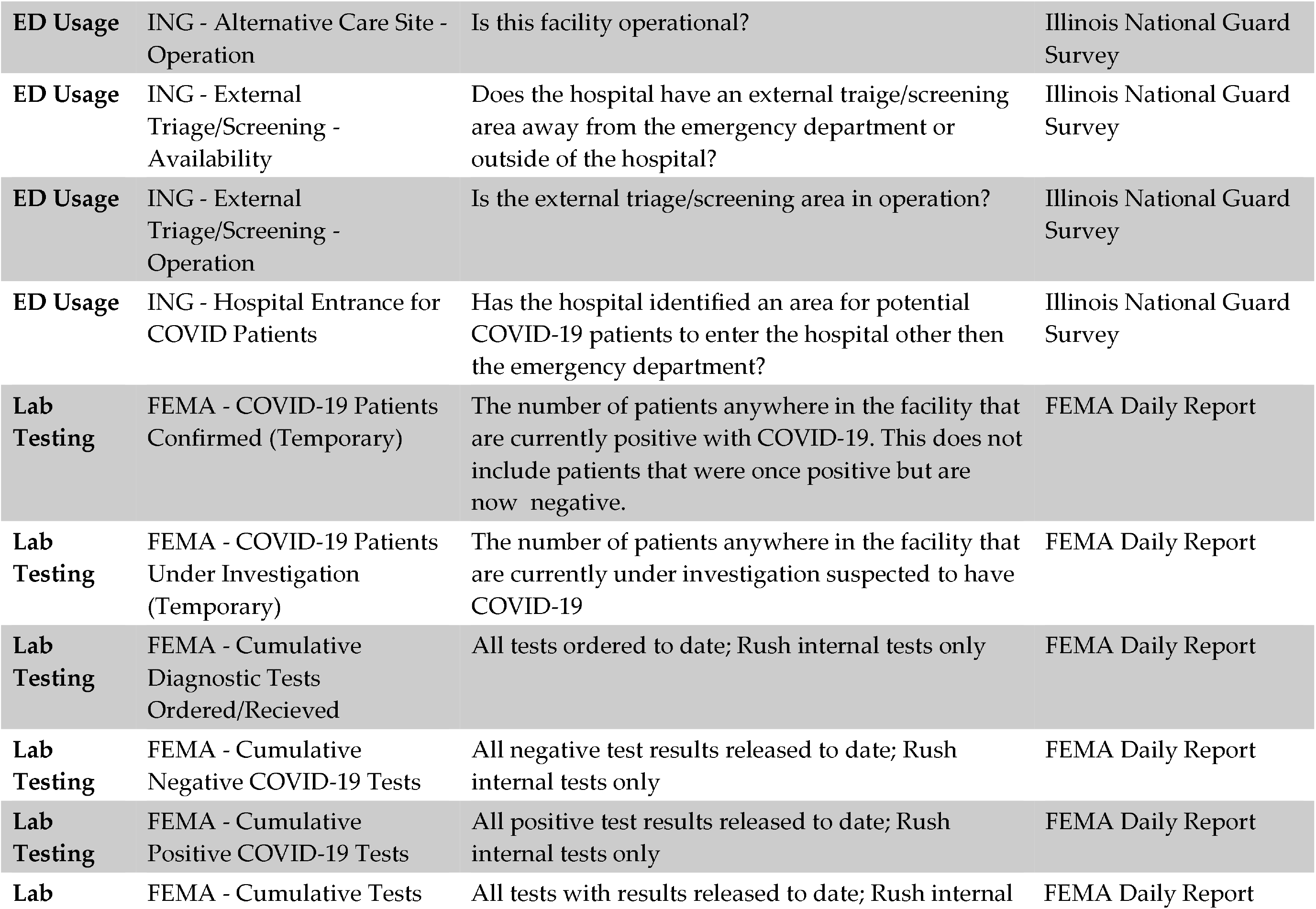
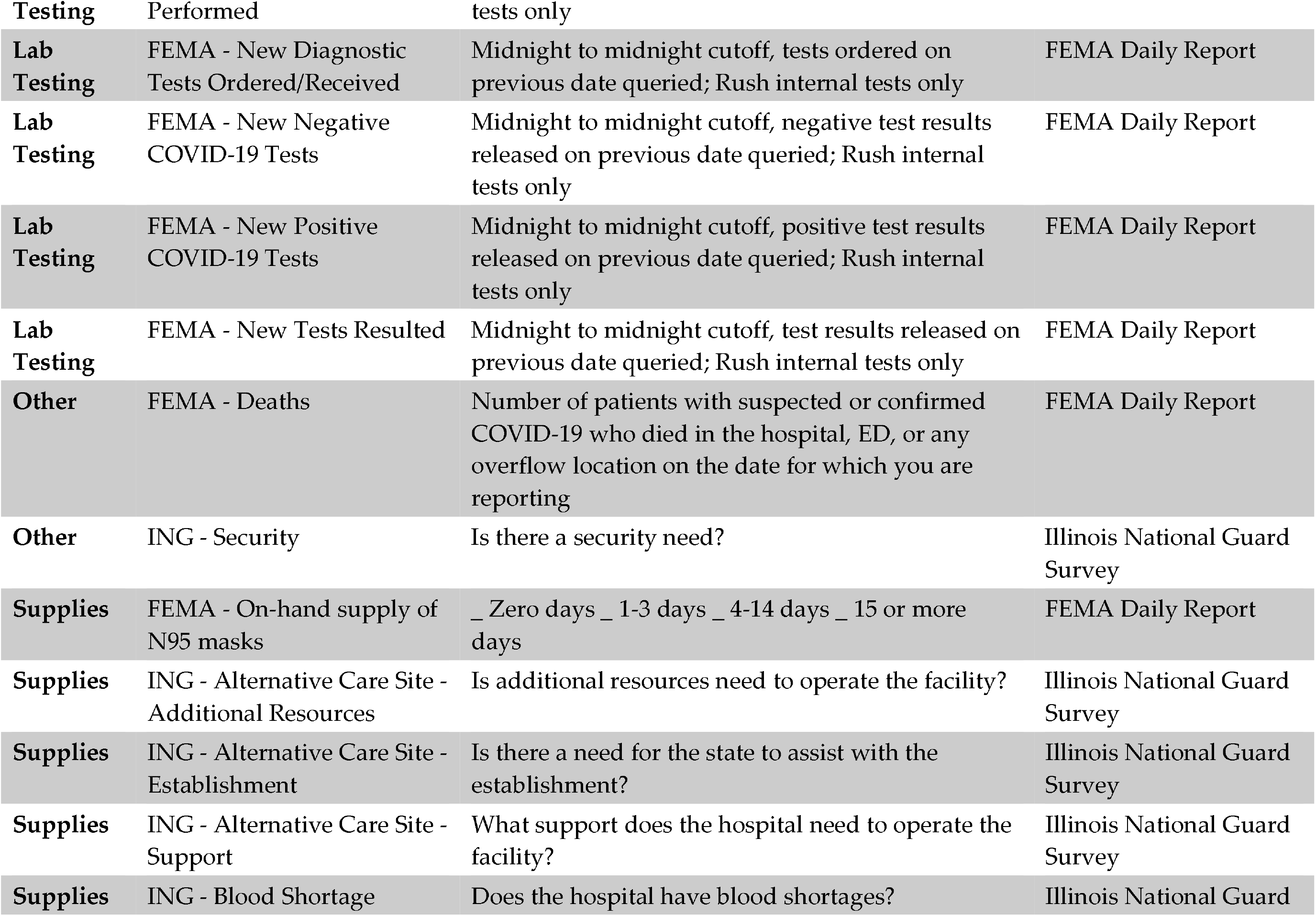
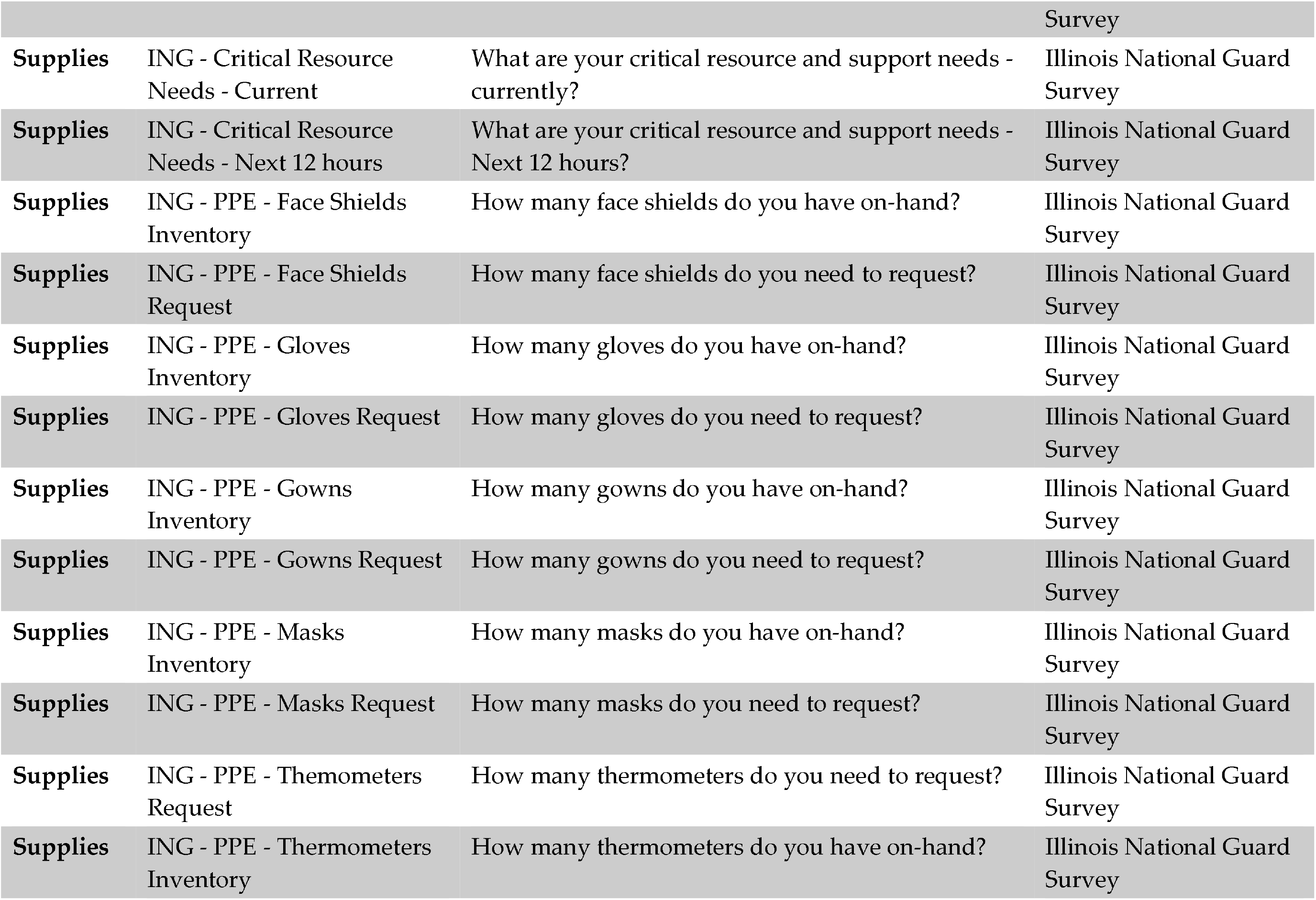
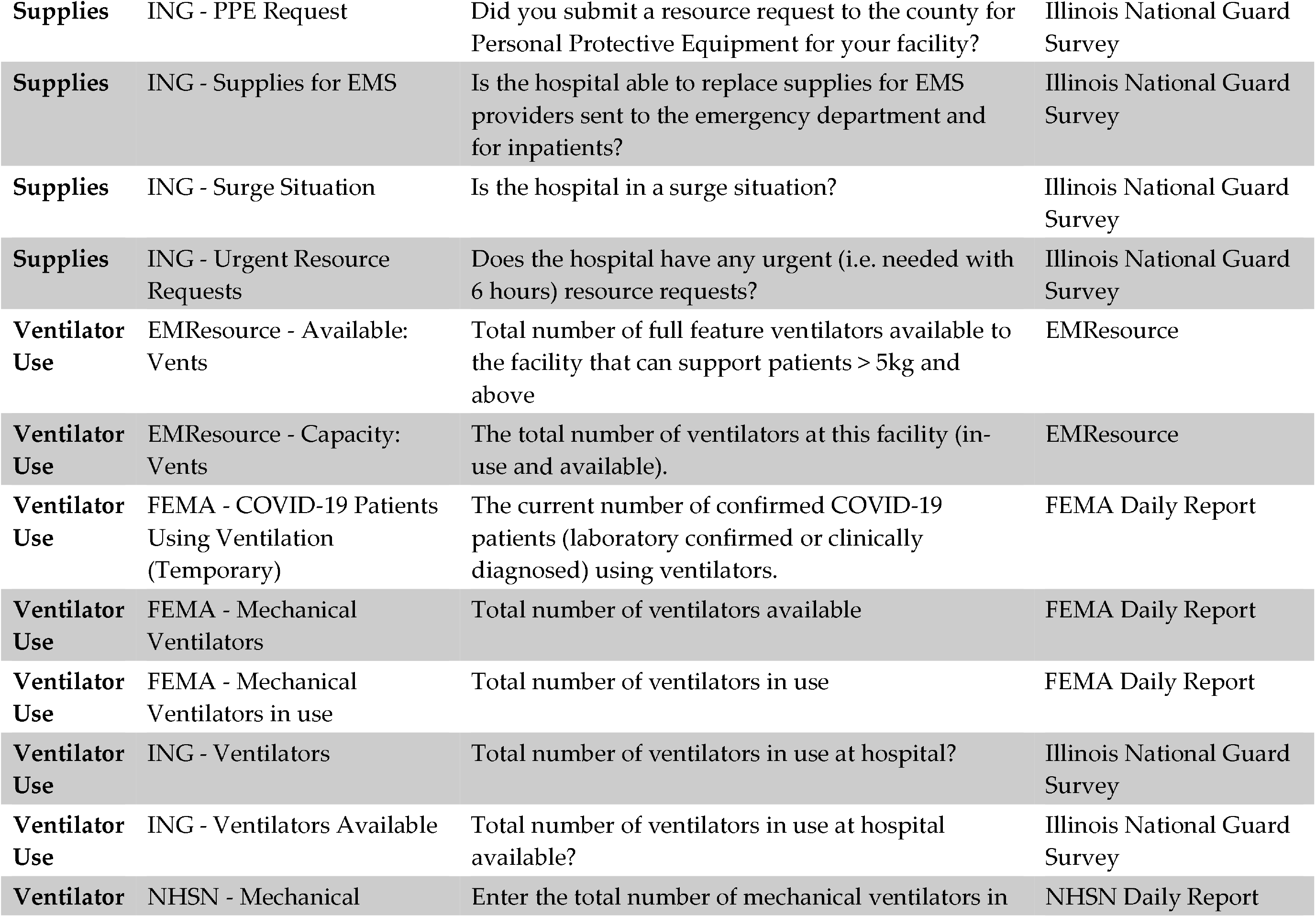
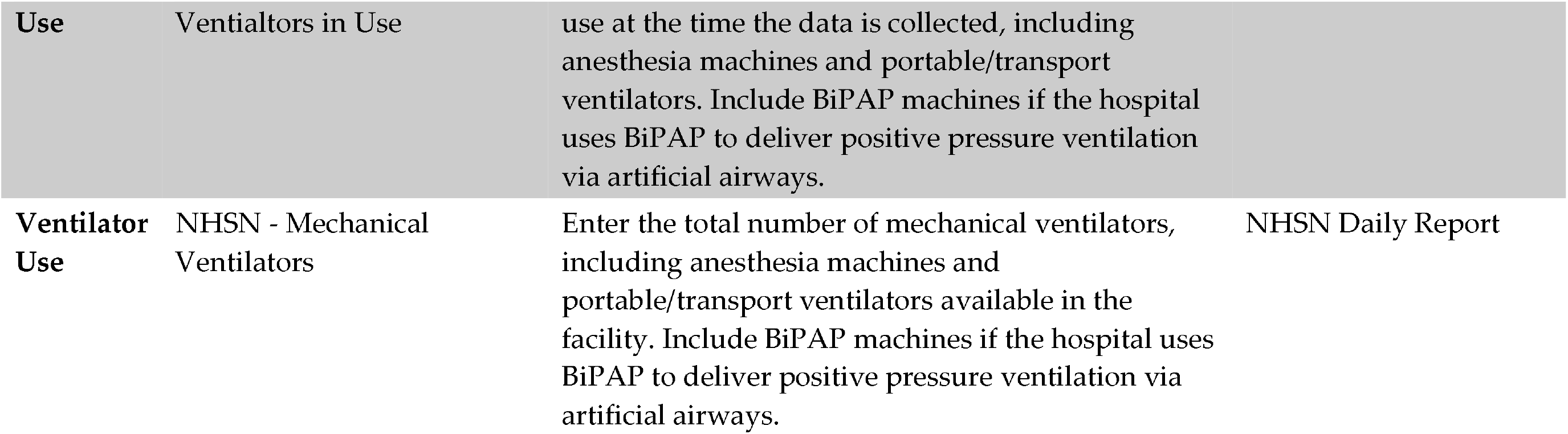
List of measures and agencies with mandated reporting in April 2020 for COVID-19 in Chicago

In response to these challenges the Chicago Department of Public Health issued a public health order requiring electronic data sharing,, and partnered with Rush University Medical Center to leverage existing HIT infrastructure for COVID-19 disease to develop a platform for data exchange. Our goals were to use regulated interoperability standards already in production to generate regional bed capacity awareness, enhance the capture of epidemiological risk factors and clinical variables among COVID-19 tested patients, and reduce the administrative burden of reporting for stakeholders in a manner that could be replicated by other public health agencies. As an example of clinically relevant fields of interest for reporting, we compared available fields in data feeds to the CDC PUI form. We also evaluated the completeness of various data sources supplied to the platform and the capacity to link these sources.

## Methods

### Setting

Chicago is the third largest city in the country and has witnessed a high disease burden of COVID-19, with over 56,000 lab confirmed cases among Chicago residents as of July 15, 2020. This project was conducted by CPDH in partnership with Rush University Medical Center, which was made a third-party agent of CDPH to develop and support the analytics and provide the infrastructure to support the data collection.

### Public health notice

On April 6, 2020, the CDPH issued public health order 2020-4 requiring hospitals in Chicago to share electronic health record data with the public health department(9) for all patients tested for COVID-19. The order outlined a constrained set of data to be submitted for all COVID-19 tested patients. This order was disseminated through CDPH’s clinical health alert network, posted on the department’s website, and shared with city hospital leadership on calls. CDPH constituted a governance committee comprised of medical directors and informaticists from hospital systems in Chicago.

### Data Feeds

Electronic Lab Records (ELR) feeds were accessed from the Illinois’ National Electronic Disease Surveillance System (I-NEDSS) to provide baseline information on lab confirmed cases in the city. To meet public health order 2020-4, Chicago hospitals were provided multiple mechanisms to submit consolidated clinical data architecture (CCDA) records for COVID-19 tested patients. This included: a) via a secure mailbox that used the DIRECT protocol, a recognized data standard by the Office of the National Coordinator, for the one-way transmission of electronic health records from, as an example, to a centralized instance of Epic for the city, or b) directly to CDPH’s Azure instance via DIRECT or an API, which could ingest the CCDA records. In either case the CCD As were parsed into a database within a dedicated tenant in Azure for analytics. Additionally, a third dataset of NHSN patient safety and hospital capacity was included, which hospitals were asked to either enter into a REDCap database or send electronically to the Azure tenant.

### Technical Evaluation

At the project start, we developed the requirements of a solution to collect data from sites and produce the required analytics. We evaluated the gap between the existing CDC PUI form fields and the electronic data elements available in federal standards based data feeds, and developed a crosswalk of reporting requirements to ensure that the data set could function as a reporting gateway for sites and reduce the burden of reporting. Feeds evaluated were electronic laboratory reporting, consolidated clinical data architecture, and Fast Healthcare Interoperability Resources fields. Comparisons were made based on the latest specifications documents and potential field presence for each. Missingness and usefulness were evaluated among CCDA and ELR feeds. Missingness refers to the presence of data in the field. Usefulness refers to clean, complete information in data field; <100% indicates “unknown” (race, ethnicity, address, etc); for address (PO Boxes, Unknown, Homeless, N/A); phone (no phone, bad number/not enough numbers); ZIP codes <5 digits or 99999 or 00000 or UUUUU etc. Records were deduplicated using name and date of birth. Record match rate between CCDA and ELR data feeds was assessed using last and first name and date of birth.

This investigation was part of the ongoing public health response to COVID-19; thus, was determined to be non-research, public health surveillance and exempt from human subjects’ research regulations.

## Results

### State surveillance system baseline reporting

In Chicago, a significant proportion of reported cases of SARS CoV-2 infection are reported through ELR. As of June 30, 2020, ELR alone provided 73.7% of cases, while ELR combined with other modalities accounted for 94% of reported cases. ELR data reports key fields requested in the CDC PUI form (Table 2) but not all; fields routinely absent from ELR feeds included travel histories, clinical symptoms, and comorbidities.

**Table 2.**
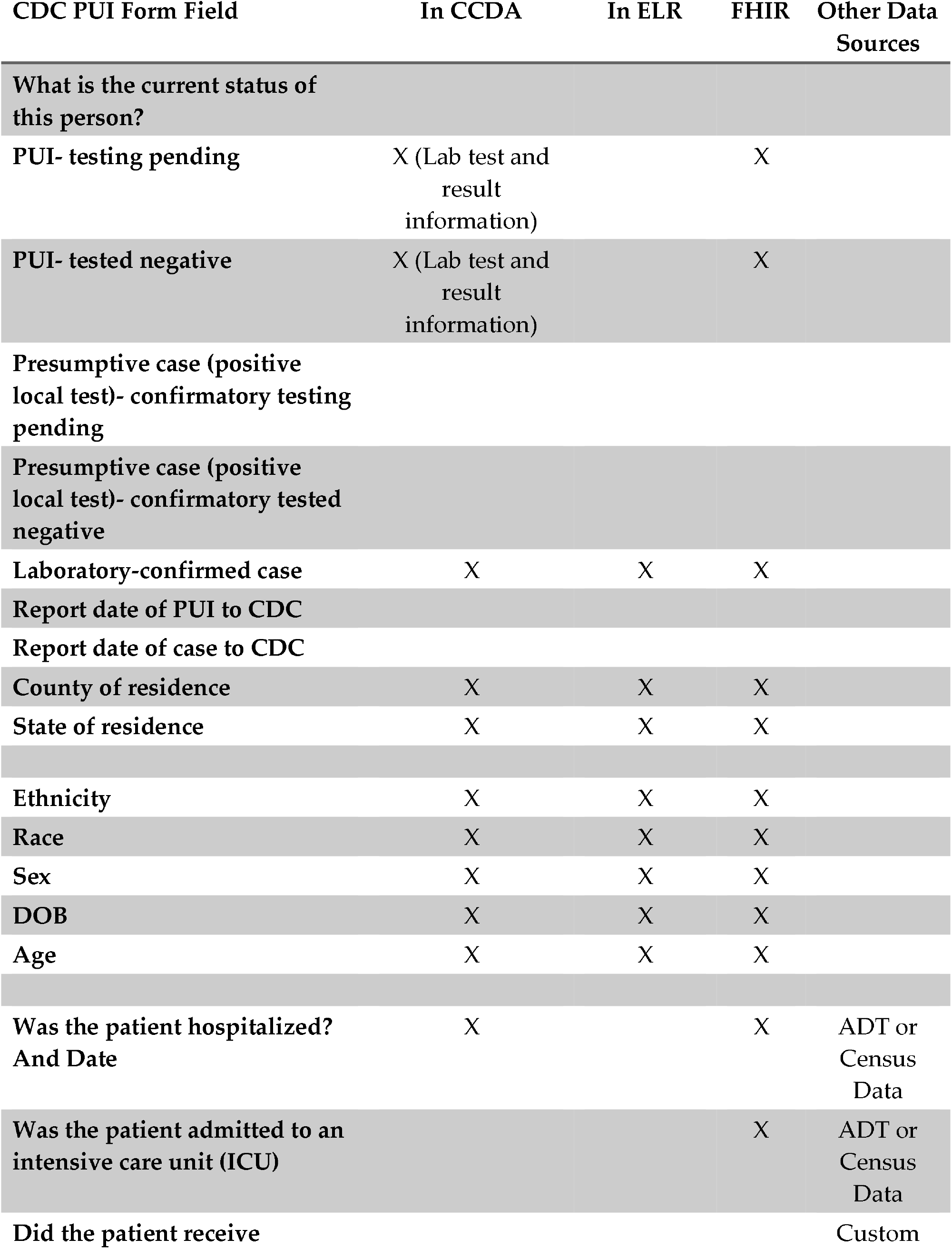
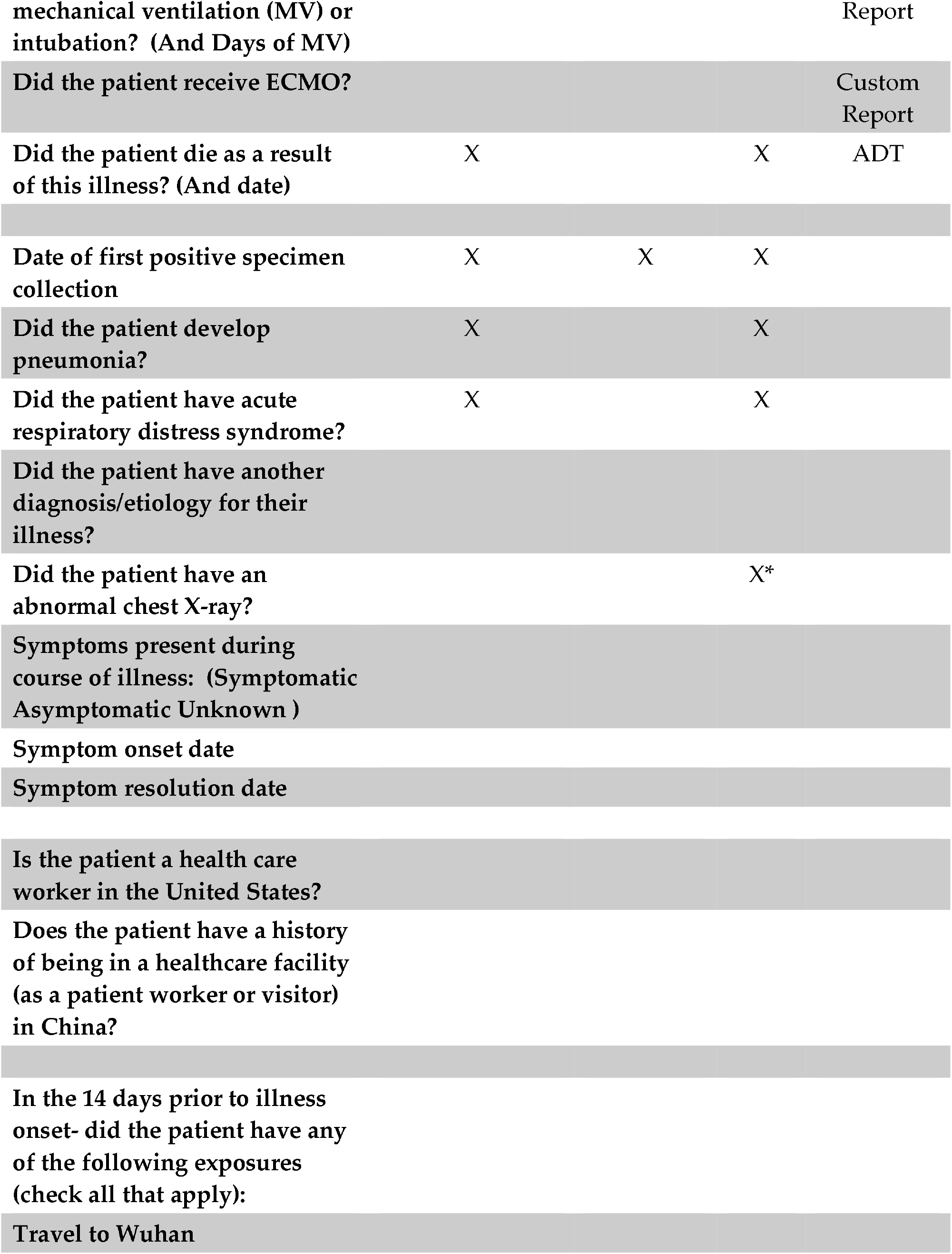
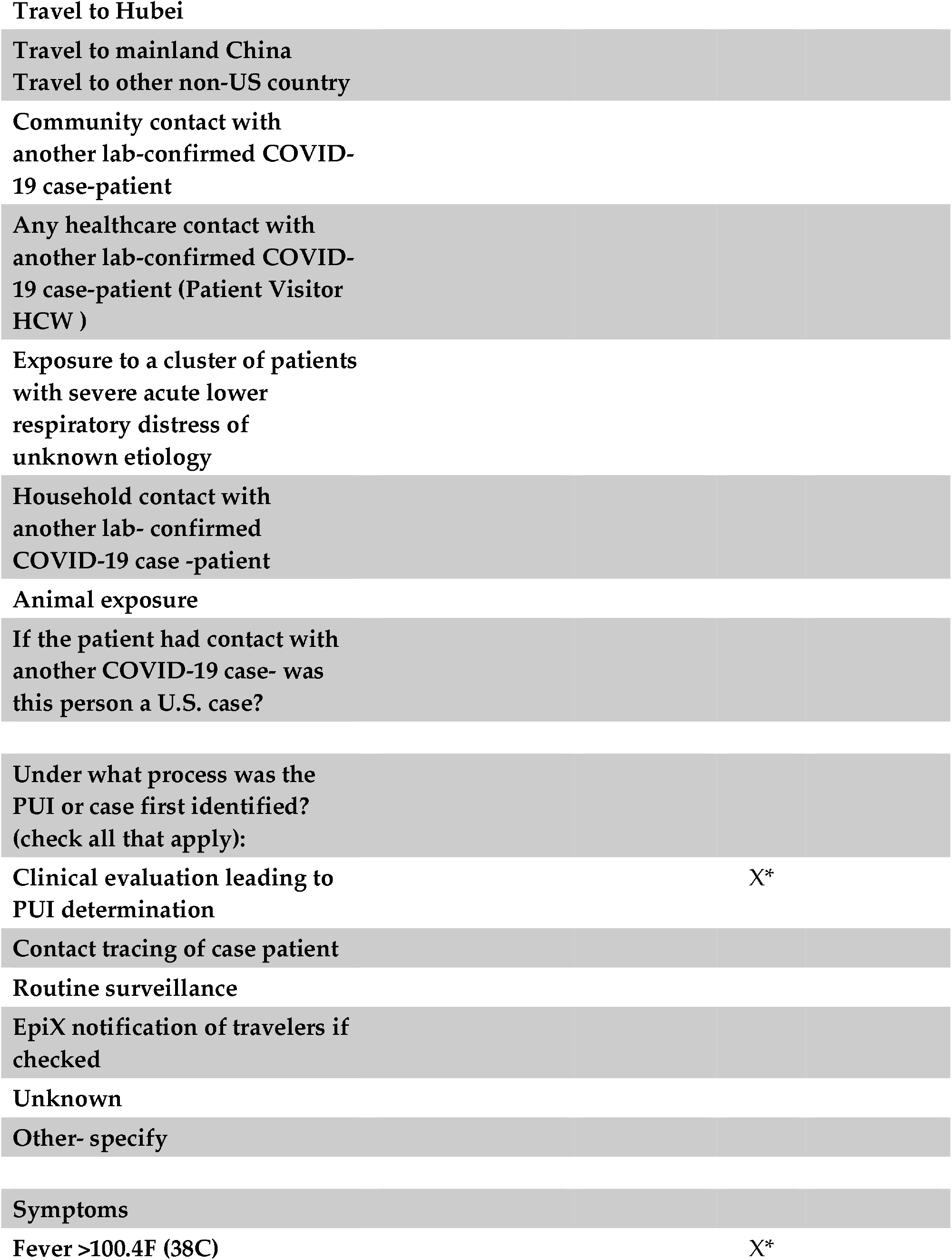
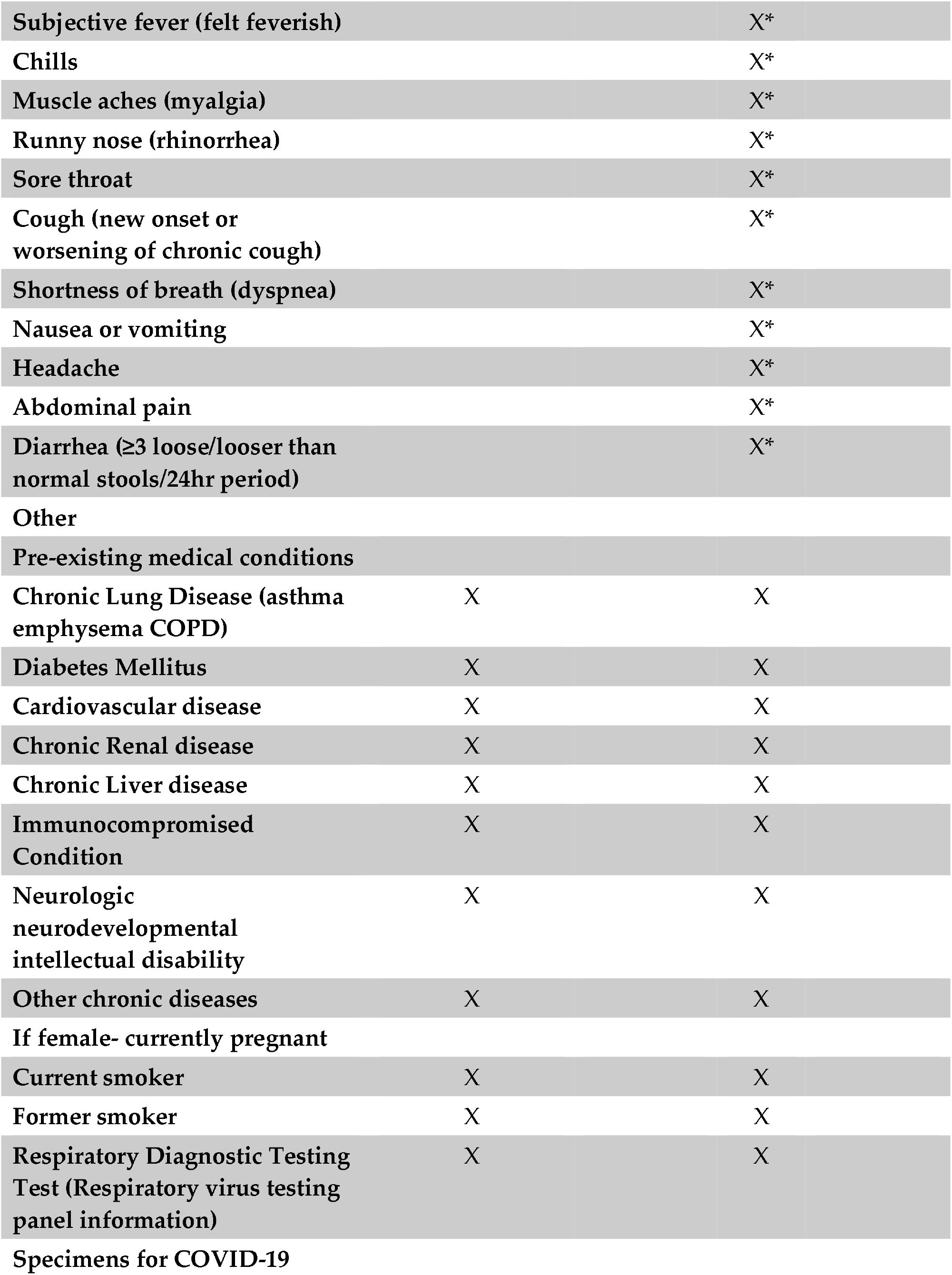
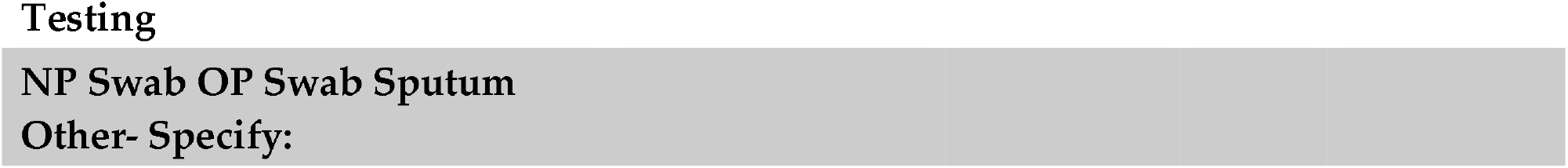
Crosswalk table to compare coverage of PUI form fields and ELR, CCDA, and FHIR Resources *If Notes are shared through FHIR Resources

### Response to public health notice

A public health order was shared via the Health Alert Network in Chicago to all eligible institutions. The order mandated the sharing with CDPH of three main data types: 1) ELR feeds of SARS-CoV-2 tested individuals, which were an existing state mandate; 2) CCDA records from hospitals for SARS-CoV-2 tested patients; and 3) NHSN capacity module reporting, which was asked to be sent centrally to CDPH. These data were requested to be sent at a minimum once per day, by 10 am. Sites were also provided contact information for key Rush University Medical Center personnel who were leading the implementation. A series of calls with hospital technical staff were conducted by the Rush CIO (SR) to introduce the project, review rationale, and describe technical approaches.

An Azure hosted and isolated environment was established, with five individual modalities for connectivity; all feeding into a centralized data hub from more than 40 organizations and hundreds of thousands of transactions per week. Over the next 30 days, sites were approached for data sharing; a CDPH data governance committee composed of chief medical officers and chief medical informatics officers from select institutions was created through which issues could be discussed and additional roadmaps could be generated; collaboration with Epic and Cerner EHR developers was established, and mechanisms for enterprise scale sharing was created; and data was sent centrally to the CDPH azure instance.

### Technical Architecture

An overview of the technical architecture of the project is shown in Figure 1, and was designed to maximize security and privacy of data, keeping the CDPH at the center of data use. At a high level, because of the tools from meaningful use adoption, connections existed between stakeholders in the system, and could support secure file sharing with the ability to choose records based on criteria. These tools included: a) standards based representation of clinical data (e.g. CCDA) b) secure methods of data transport both within and external to EHR systems (e.g. CareEverywhere within Epic, Direct mailboxes, and API based authenticated pathways) and c) and existing implementation of complex public health rules within EHRs to identify cases, and submit to public health (e.g. ELR reporting).

**Figure 1.**
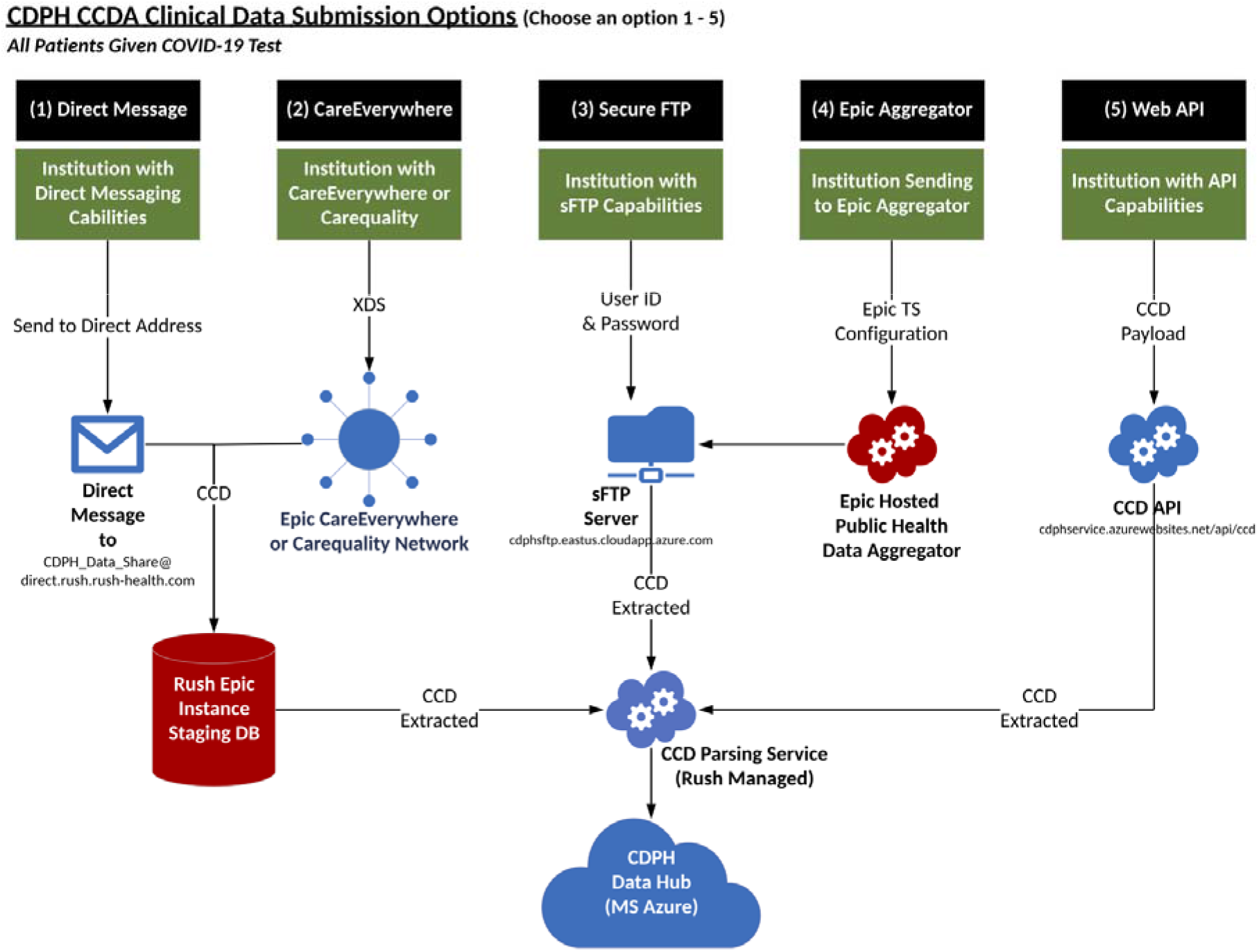
High level architecture of CDPH Data Hub

A cloud-based environment was created that was totally isolated from the Rush EHR instance and Rush patient records. This environment was built to support over 40 organizations within the city of Chicago and designed to scale across public health departments.

ELR data feeds were the most straightforward to use in the model, as existing connections between hospital systems were present for communicable disease reporting. Hospitals were required to implement new logic at the outset of COVID-19 infections in Chicago to identify and report lab identified cases of COVID-19 to public health, and tested patients as those are PUIs. ELR feeds are submitted to the state public health agency, which makes these available to the CDPH/local health departments.

To isolate data, Rush created an isolated Azure Data Repository including SQL Warehouse and a CosmosDB for survey forms data was created. We found that not all XDS and direct messages could avoid our EHR instance in our plan, which had to be addressed to enforce separation of data. We addressed this by pulling data from the the Epic staging area. In addition, infrastructure components were created that included an XDS service server, Direct Message communication, a CCD to FHIR service, and integration with Epic via a community health aggregator. Apigee handled the API layer, and services were handled behind Apigee for token control. Data collection via manual entries were handled via REDCap forms with integration via the API into the Azure environment.

### Governance

Data governance was planned from the project beginning to aid in consensus and principles for data use. While the local health department, with its public health orders, was a necessary recipient and user of the data, participants recognized the value of a larger sharing initiative, plus site participation to engage on use cases and mechanisms to leverage the information. The committee was composed of 12 site CMO, CMIO, or technical leads. These leaders also brought content and guidance back to site participants, and sought to bridge varying degrees of internal technical capabilities among systems. The committee met weekly and helped to build trust among participating sites. General principles were modeled after rules implemented for use of CMS data(10) and were established among sites through this committee. These were:

*Openness:* Promoting and facilitating the open sharing of knowledge about COVID-19 data.

*Communication:* Promoting partnerships across the region to eliminate duplication of effort, a source of truth for regional data that may enable reduced administrative burden, and a valuable regional and national resource.

*Accountability:* Ensuring compliance with approved data management principles and policies. Understanding the objectives of current and future strategic or programmatic initiatives and how they impact, or are impacted by, existing data management principles and policies as well as current privacy and security protocols.

### Reporting of bed, supply, and clinical capacity

Metrics mandated for reporting to multiple agencies and groups for Chicago hospitals at the time of the hub creation are shown in Table 1. In this inventory, over 100 measures to 4 systems were required: the National Healthcare Safety Network, EMResource, FEMA, and the Illinois National Guard. The systems measure Bed usage, ED Usage, Ventilator usage, Supply usage and need, and Lab testing. Of note, 57 different bed usage measures alone exist between the 4 systems. Though metrics shown had similar definitions, these still require separate administrative efforts for the collection and reporting of the data.

As of July 31, 2020, 14 hospitals in Chicago were reporting data to the hub. For bed capacity reporting, 7 were reporting NHSN data through manual data submission, 2 were reporting through electronic queries from their EHR with electronic submission to the hub, and 14 were submitting to EMResource.

### Completeness of reporting via ELR and CCDA

88,906 total persons from CCDA data among 14 facilities, and 408,741 persons from ELR records among 88 facilities, were submitted. Table 3 shows the volume and completeness of data feeds related to COVID-19 as obtained from CCDA data and with ELR feeds. Individuals with records in these feeds were those with diagnostic testing (i.e molecular) with a Chicago address through July 31, 2020. For those individuals with more than one test reported, data have been deduplicated. Among individuals with CCDA records submitted, 13.1%(11,657) had positive tests, compared with 13.2% (53,968) among ELR feeds.

**Table 3.**
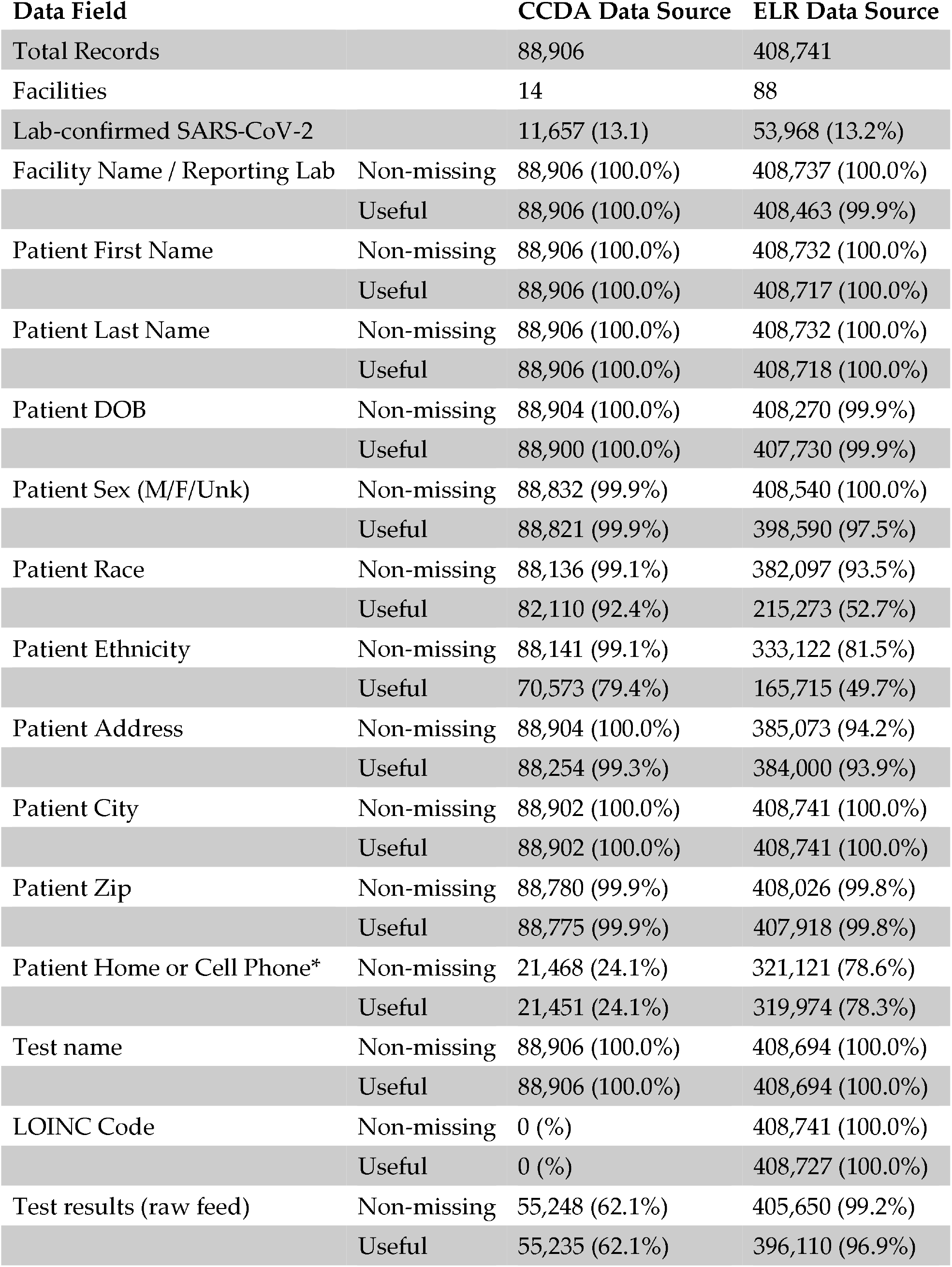
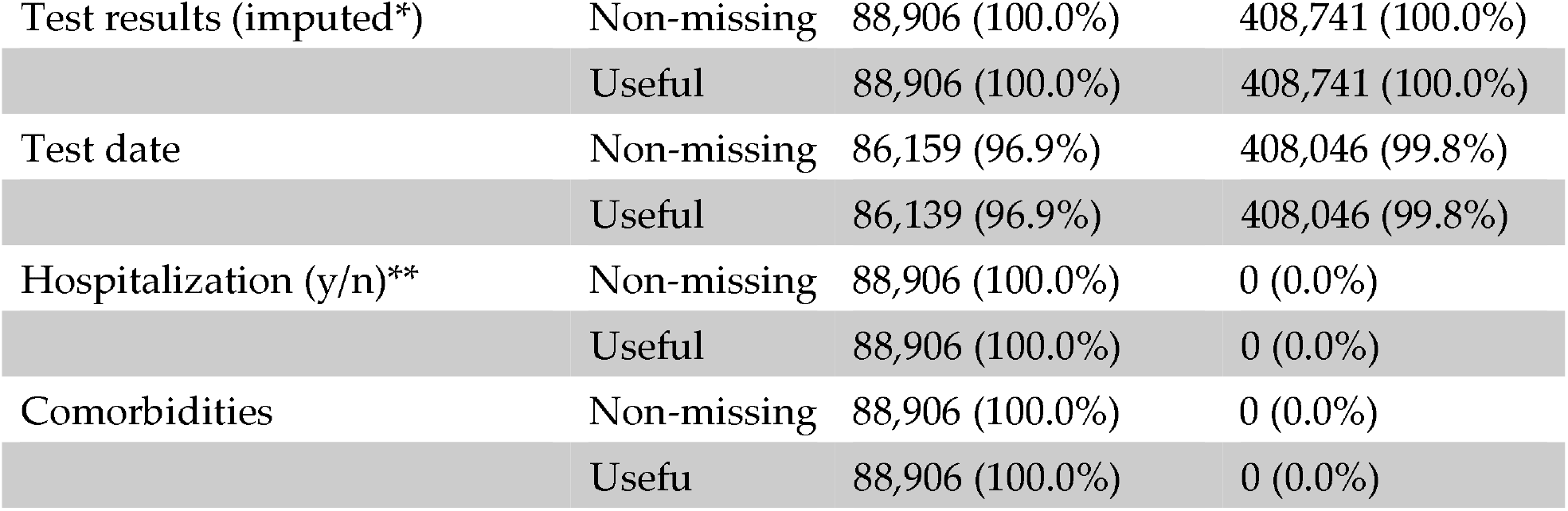
Completeness of data submitted via CCDA and ELR *Cell and Home phone not differentiated in ELR feed; Non-missing refers to populated data field; Useful refers to clean, complete information in data field; <100% indicates “unknown” (race, ethnicity, address, etc); for address (PO Boxes, Unknown, Homeless, N/A); phone (no phone, bad number/not enough numbers); ZIP codes <5 digits or 99999 or 00000 or UUUUU etc.

CCDA data provided an improvement in the quality of data available for surveillance. ELR feeds had gaps in the usability, or quality, of race and ethnicity data. With the addition of CCDA information, this content improved. CCDA was also highly complete with <5% for all records types with the exception of patient cell phone, though a contact phone number was highly complete. CCDA, though covering fewer records, also had information related to encounters and hospitalization as well as the presence of comorbidities.

CCDA and ELR data feeds were matched by name and date of birth among 90.1% of patients. With matching, some improvement in data completeness for the three most incomplete fields was noted: race completeness improved to 20.6% (1.4% improvement), ethnicity improved to 41.5% (3.5% improvement), and phone number improved to 31.4%(2.8% improvement)

For presentation, data were displayed on a dashboard available for CDPH analysts, via the Azure Power BI platform and are shown in Figure 2.

**Figure 2.**
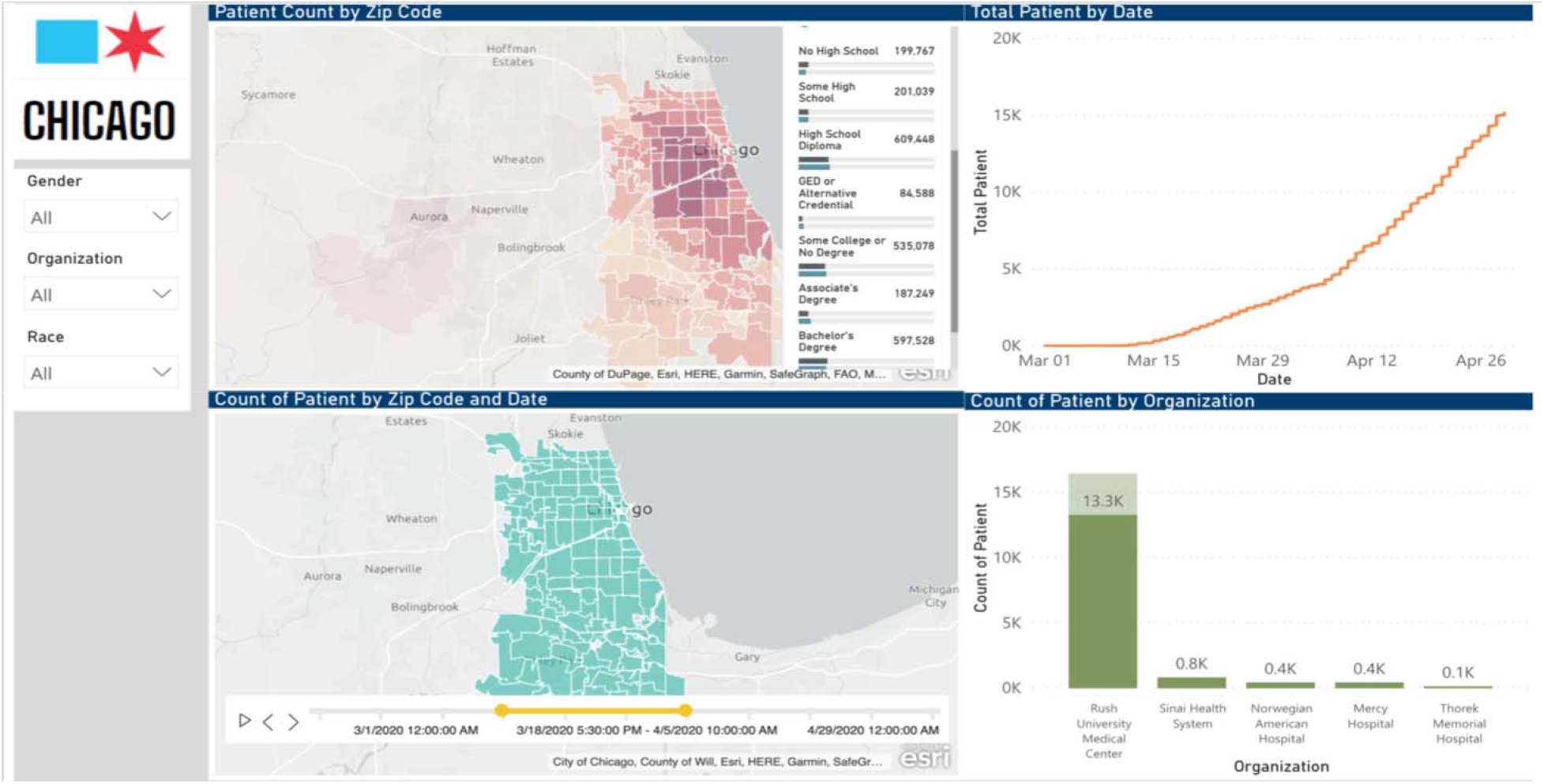
Epidemiologic dashboards for assessment of outbreak, CDPH data hub

## Discussion

In this report, we describe the development of a city-wide public health data hub for the surveillance of COVID-19 infection. We were able to assess the completeness of existing ELR feeds, augment these feeds with CCDA documents, establish secure transfer methods for data exchange, develop cloud-based architecture to enable secure data storage and analytics, and produced meaningful dashboards for the monitoring of capacity and disease burden.

Electronic laboratory reporting, or the submission electronically of positive lab tests to public health through implementation of business logic for detection, has been found in multiple studies to improve the timeliness and completeness of reporting(11–15), at potentially lower cost(16). A review prior to widespread electronic reporting use found that despite legal mandates for reporting, passive surveillance yielded completeness rates of 23-81% for communicable diseases with higher rates for active surveillance(17), and for timeliness of reporting, between 10-13 days after lab result dates(18). Electronic laboratory reporting (ELR) systems have resulted in improvements in the reporting of data to public health for surveillance. In states that have implemented ELR, the volume and timeliness have improved 2.3-4.4 fold and 3.8-7.9 days earlier, respectively(19). ELR has been a major advance in that it can improve the completeness of reporting over what is found through passive surveillance(16,20). ELR based systems use a trigger of either a test having been performed or resulting as positive to start a submission workflow.

However, we found that ELR data can be incomplete. In prior reports, ELR data has been found to vary in its completeness: the completeness of fields reported via ELR within basic HL7 v 2.x messages ranges from 38% (race) to 98% (date of birth)(20). To improve completeness, groups have proposed 1) increased mandatory fields in ELR HL7 2.x messages(19); 2) augmenting ELR feeds with data from a health information exchange, which improved completeness for race to 60% (20); and 3) electronic case report forms which are completed either through automated data capture or manual completion(21). Significant limitations in case reporting have identified during the COVID-19 pandemic, including limited data on key variables such as age, race/ethnicity, hospitalization, and ICU status(22).

Laboratory reporting does not provide all of the information needed for adequate case investigation, however. As our data shows, demographic and risk factor information may not be complete in the HL7 feeds for ELR, and case report forms continue to have a critical role in the work of public health practice. Additionally, co-morbid conditions, a significant predictor of disease outcome, are not captured. We found that CCDA data had a broader set of clinical fields, and have the advantage of providing valuable comorbidity information. While only small improvements in completeness were achieved, a high match rate to ELR data makes CCDA a compelling addition to ELR to improve the analytic power of public health data sets.

Initiatives to standardize and automate case report form completion have been developed (23) and piloted (24), which have shown promise at reducing the time to complete reporting. Similar to our results, others have found that health information exchanges show value in prepopulating key elements for reporting through automated matching and searches in the patient record(25). The use of FHIR(26) may provide a viable path for automatic of public health case reporting and reduce administrative burden: when combined with an ELR based trigger for a case (in this example, sexually transmitted infection cases), an app that executed a FHIR-based query could complete an electronic case report form in 85% of cases(21). Additionally, all the key components of FHIR based workflows for public health reporting are often in place(27).

A feature of our solution is that it supports the central role of local health departments in data aggregation and reporting. An important component of the public health response in many communities is “home rule” for public health agencies(28), or local jurisdiction and control of policy and approach for local health departments. Present in 48 states, home-rule law empowers local governments to address public health issues and fill gaps in the patchwork of national and state based public health response. In the current pandemic, robust local responses that can enable targeted interventions and planning can allow more sophisticated preparedness planning, pandemic control, and epidemiological analysis.

For the most efficient exchange of data, standards for the structure of data shared, and on the semantic representation of information are critical. In this context, the technical and non-technical handshakes and handoffs related to data are key factors in successful programs. In this setting, technical handshakes are the trust relationships between systems to enable data sharing: the ability to use both authenticated API based transfers, and Direct Mailbox shares accelerated time to implementation for the project. Technical handoffs were the ability to have seamless data parsing because of robust standards implemented via meaningful use. The ability to leverage CCDA to increase the completeness of overall PUI reporting, given the greater coverage of fields in the PUI form by CCDA files, is a sign of the value of federal standards for clinical data interchange.

Of more importance were the non-technical “handshakes” - i.e. relationship building and the development of consensus among institutions to enable sharing of data - and “handoffs” - the partnerships between public and private entities. A data governance committee was essential to promote trust, enabled the scaling of the program to new data sets and deeper information within sets. At a time of a surge in COVID-19 cases, a private, academic partner (Rush University Medical Center) with the technical capacity was able to rapidly implement a solution.

We see this public health and clinical data registry as an informative example of the power of common standards across electronic records, and a potential template for future extension of the use of standards to improve public health surveillance.

## Data Availability

The data used for this study was reported to the health department and is not publicly available at this time.

## Acknowledgements

None

## References

1. Zhu N, Zhang D, Wang W, Li X, Yang B, Song J, et al. A Novel Coronavirus from Patients with Pneumonia in China, 2019. New England Journal of Medicine [Internet]. 2020 Feb [cited 2020 Apr 27];382(8):727–33. Available from: https://doi.org/10.1056/NEJMoa2001017

2. WHO Director-General’s opening remarks at the media briefing on COVID-19 -11 March 2020 [Internet]. [cited 2020 Apr 27], Available from: https://www.who.int/dg/speeches/detail/who-director-general-s-opening-remarks-at-the-media-briefing-on-covid-19---11-march-2020

3. Syndromic Surveillance (SS) | Meaningful Use | CDC [Internet]. 2020 [cited 2020 Apr 25]. Available from: https://www.cdc.gov/ehrmeaningfuluse/Syndromic.html

4. Electronic Laboratory Reporting (ELR) | Meaningful Use | CDC [Internet]. 2020 [cited 2020 Apr 25]. Available from: https://www.cdc.gov/ehrmeaningfuluse/elr.html

5. Guides | Meaningful Use | CDC [Internet]. 2020 [cited 2020 Apr 25]. Available from: https://www.cdc.gov/ehrmeaningfuluse/guides.html

6. Arnold KE, Thompson ND. Building Data Quality and Confidence in Data Reported to the National Healthcare Safety Network. Infection Control & Hospital Epidemiology [Internet]. 2012 May [cited 2020 May 3];33(5):446–8. Available from: https://www.cambridge.org/core/journals/infection-control-and-hospital-epidemiology/article/building-data-quality-and-confidence-in-data-reported-to-the-national-healthcare-safety-network/DA0BlFE51A558BB4C5A4AB7CD77BE730

7. Holmgren AJ, Apathy NC, Adler-Milstein J. Barriers to hospital electronic public health reporting and implications for the COVID-19 pandemic. Journal of the American Medical Informatics Association [Internet]. [cited 2020 Jul 17]; Available from: https://academic.oup.com/jamia/article/doi/10.1093/jamia/ocaall2/5842141

8. Strengthening the Public Health Infrastructure: The Role of Data in Controlling the Spread of COVID-19 [Internet]. 2020. Available from: http://www.adhocresponsegroup.org/OPCAST_Public_Health_Data_Report_07-28-20.pdf

9. COVID-19 Orders [Internet], [cited 2020 May 3]. Available from: https://www.chicago.gov/content/city/en/sites/covid-19/home/health-orders.html

10. CMS Information Systems Security and Privacy Policy [Internet]. 2019. Available from: https://www.cms.gov/Research-Statistics-Data-and-Systems/CMS-Information-Technology/InformationSecurity/Downloads/CMS-IS2P2.pdf

11. Centers for Disease Control and Prevention (CDC). Automated detection and reporting of notifiable diseases using electronic medical records versus passive surveillance–massachusetts, June 2006-July 2007. MMWR Morbidity and mortality weekly report. 2008 Apr;57(14):373–6.

12. Centers for Disease Control and Prevention (CDC). Potential effects of electronic laboratory reporting on improving timeliness of infectious disease notification–Florida, 2002-2006. MMWR Morbidity and mortality weekly report. 2008 Dec;57(49): 1325–8.

13. Centers for Disease Control and Prevention (CDC). Effect of electronic laboratory reporting on the burden of lyme disease surveillance–New Jersey, 2001-2006. MMWR Morbidity and mortality weekly report. 2008 Jan;57(2):42–5.

14. Nguyen TQ, Thorpe L, Makki HA, Mostashari F. Benefits and Barriers to Electronic Laboratory Results Reporting for Notifiable Diseases: The New York City Department of Health and Mental Hygiene Experience. American Journal of Public Health [Internet]. 2007 Apr [cited 2020 Apr 26];97(Suppl 1):S142–5. Available from: https://www.ncbi.nlm.nih.gov/pmc/articles/PMC1854985/

15. Swaan C, van den Broek A, Kretzschmar M, Richardus JH. Timeliness of notification systems for infectious diseases: A systematic literature review. PloS One. 2018;13(6):e0198845.

16. Samoff E, Dibiase L, Fangman MT, Fleischauer AT, Waller AE, MacDonald PDM. We can have it all: Improved surveillance outcomes and decreased personnel costs associated with electronic reportable disease surveillance, North Carolina, 2010. American Journal of Public Health. 2013 Dec;103(12):2292–7.

17. Doyle TJ, Glynn MK, Groseclose SL. Completeness of notifiable infectious disease reporting in the United States: An analytical literature review. American Journal of Epidemiology. 2002 May;155(9):866–74.

18. Jajosky RA, Groseclose SL. Evaluation of reporting timeliness of public health surveillance systems for infectious diseases. BMC Public Health [Internet]. 2004 Jul [cited 2020 Apr 26];4:29. Available from: https://www.ncbi.nlm.nih.gov/pmc/articles/PMC509250/

19. Rajeev D, Staes CJ, Evans RS, Mottice S, Rolfs R, Samore MH, et al. Development of an electronic public health case report using HL7 v2.5 to meet public health needs. Journal of the American Medical Informatics Association: JAMIA. 17(1):34–41.

20. Dixon BE, McGowan JJ, Grannis SJ. Electronic laboratory data quality and the value of a health information exchange to support public health reporting processes. AMIA Annual Symposium proceedings AMIA Symposium. 2011;2011:322–30.

21. Dixon BE, Taylor DE, Choi M, Riley M, Schneider T, Duke J. Integration of FHIR to Facilitate Electronic Case Reporting: Results from a Pilot Study. Studies in Health Technology and Informatics. 2019 Aug;264:940–4.

22. CDCMMWR. Severe Outcomes Among Patients with Coronavirus Disease 2019 (COVID-19) United States, February 12March 16, 2020. MMWR Morbidity and Mortality Weekly Report [Internet]. 2020 [cited 2020 Jun 5];69. Available from: https://www.cdc.gov/mmwr/volumes/69/wr/mm6912e2.htm

23. Mac Kenzie WR, Davidson AJ, Wiesenthal A, Engel JP, Turner K, Conn L, et al. The Promise of Electronic Case Reporting. Public Health Reports [Internet]. 2016 Nov [cited 2020 Apr 26];131(6):742-6. Available from: https://doi.org/10.1177/0033354916670871

24. Whipple A, Jackson J, Ridderhoff J, Nakashima AK. Piloting Electronic Case Reporting for Improved Surveillance of Sexually Transmitted Diseases in Utah. Online Journal of Public Health Informatics [Internet]. 2019 Sep [cited 2020 Apr 25];11(2). Available from: https://www.ncbi.nlm.nih.gov/pmc/articles/PMC6788887/

25. Painter I, Revere D, Gibson PJ, Baseman J. Leveraging public health’s participation in a Health Information Exchange to improve communicable disease reporting. Online Journal of Public Health Informatics. 2017;9(2):e186.

26. Index - FHIR v4.0.1 [Internet], [cited 2020 May 2], Available from: https://www.hl7.org/fhir/

27. Mishra NK, Duke J, Lenert L, Karki S. Public health reporting and outbreak response: Synergies with evolving clinical standards for interoperability. Journal of the American Medical Informatics Association [Internet]. [cited 2020 Jul 17]; Available from: https://academic.oup.com/jamia/article/doi/10.1093/jamia/ocaa059/5848746

28. McCarty KL, Nelson GD, Hodge JG, Gebbie KM. Major Components and Themes of Local Public Health Laws in Select U.S. Jurisdictions. Public Health Reports [Internet]. 2009 [cited 2020 Jul 16];124(3):458–62. Available from: https://www.ncbi.nlm.nih.gov/pmc/articles/PMC2663884/

